# Time course of phosphorylated tau181 in blood across the Alzheimer’s disease spectrum

**DOI:** 10.1101/2020.07.13.20152025

**Authors:** Alexis Moscoso, Michel J. Grothe, Nicholas J. Ashton, Thomas K. Karikari, Juan Lantero Rodriguez, Anniina Snellman, Marc Suárez-Calvet, Henrik Zetterberg, Kaj Blennow, Michael Schöll, for the Alzheimer’s Disease Neuroimaging Initiative

**Affiliations:** Department of Psychiatry and Neurochemistry, Institute of Neuroscience and Physiology, The Sahlgrenska Academy, University of Gothenburg, Sweden; Wallenberg Centre for Molecular and Translational Medicine, University of Gothenburg, Sweden; Unidad de Trastornos del Movimiento, Instituto de Biomedicina de Sevilla (IBiS), Hospital Universitario Virgen del Rocío/CSIC/Universidad de Sevilla, Sevilla, Spain; King’s College London, Institute of Psychiatry, Psychology & Neuroscience, Maurice Wohl Clinical Neuroscience Institute, London, UK; NIHR Biomedical Research Centre for Mental Health & Biomedical Research Unit for Dementia at South London & Maudsley NHS Foundation, London, UK; Turku PET Centre, University of Turku, Kiinamyllynkatu 4-8, FI-20520 Turku, Finland; Barcelonaβeta Brain Research Center (BBRC), Pasqual Maragall Foundation. Barcelona, Spain; IMIM (Hospital del Mar Medical Research Institute), Barcelona, Spain; Servei de Neurologia, Hospital del Mar, Barcelona, Spain; Centro de Investigación Biomédica en Red de Fragilidad y Envejecimiento Saludable (CIBERFES), Madrid, Spain; Clinical Neurochemistry Laboratory, Sahlgrenska University Hospital, Mölndal, Sweden; Department of Neurodegenerative Disease, UCL Queen Square Institute of Neurology, University College London, London, UK; UK Dementia Research Institute at University College London, London, UK

**Keywords:** Alzheimer’s disease, blood biomarkers, tau, positron emission tomography, cerebrospinal fluid

## Abstract

Tau phosphorylated at threonine 181 (p-tau181) measured in blood plasma has recently been proposed as an accessible, scalable, and highly specific biomarker for Alzheimer’s disease. Longitudinal studies, however, investigating the temporal dynamics of this novel biomarker are lacking. It is therefore unclear when in the disease process plasma p-tau181 increases above physiological levels and how it relates to the spatiotemporal progression of Alzheimer’s disease-characteristic pathologies. We aimed to establish the natural time course of plasma p-tau181 across the sporadic Alzheimer’s disease spectrum in comparison to those of established imaging- and fluid-derived biomarkers of Alzheimer’s disease. We examined longitudinal data from a large prospective cohort of elderly individuals enrolled in the Alzheimer’s Disease Neuroimaging Initiative (ADNI) (*n*=1067) covering a wide clinical spectrum from normal cognition to dementia, and with measures of plasma p-tau181 and an [18F]florbetapir amyloid-β (Aβ) positron emission tomography (PET) scan at baseline. A subset of participants (*n*=864) also had measures of Aβ_1-42_ and p-tau181 levels in cerebrospinal fluid (CSF), and another subset (*n*=298) had undergone an [18F]flortaucipir tau PET scan six years later. We performed brain-wide analyses to investigate the associations of plasma p-tau181 baseline levels and longitudinal change with progression of regional Aβ pathology and tau burden six years later, and estimated the time course of changes in plasma p-tau181 and other Alzheimer’s disease biomarkers employing a previously developed method for the construction of long-term biomarker temporal trajectories using shorter-term longitudinal data. Spline regressions demonstrated that earliest plasma p-tau181 changes occurred even before Aβ-markers reached abnormal levels, with greater rates of change correlating with increased Aβ pathology. Voxel-wise PET analyses yielded relatively weak, yet significant, associations of plasma p-tau181 with Aβ pathology in early-accumulating brain regions in cognitively healthy individuals, while the strongest associations with Aβ were observed in late-accumulating regions in patients with mild cognitive impairment. Cross-sectional and particularly longitudinal measures of plasma p-tau181 were associated with widespread cortical tau aggregation six years later, covering temporo-parietal regions typical for neurofibrillary tangle distribution in Alzheimer’s disease. Finally, we estimated that plasma p-tau181 reaches abnormal levels approximately 6.5 and 5.7 years after CSF- and PET-measures of Aβ, respectively, following similar dynamics as CSF p-tau181. Our findings suggest that plasma p-tau181 increases are associated with the presence of widespread cortical Aβ pathology and with prospective Alzheimer’s disease-typical tau aggregation, providing clear implications for the use of this novel blood biomarker as a diagnostic and screening tool for Alzheimer’s disease.

## Introduction

Non-physiological accumulation of amyloid-β (Aβ) peptides into extracellular plaques and aggregation of hyperphosphorylated tau protein into intracellular neurofibrillary tangles (NFT) constitute the neuropathological signature of Alzheimer’s disease in the human brain (Hyman *et al*., 2012). While the reliable detection of these pathologic changes has traditionally been restricted to histopathological examination *post mortem*, current positron emission tomography (PET) and cerebrospinal fluid (CSF) biomarkers have enabled their accurate assessment *in vivo* (Blennow *et al*., 2015; Schöll *et al*., 2019). These biomarkers thus provide clinically relevant information for the detection and differential diagnosis of Alzheimer’s disease (Dubois *et al*., 2014; Ossenkoppele *et al*., 2018), its progression (Hanseeuw *et al*., 2019), as well as patient management (Rabinovici *et al*., 2019), representing key modalities for obtaining an accurate, individualized picture of a patient’s pathologic profile. However, these specialized techniques are limited by relatively high costs, invasiveness, and/or limited availability in routine clinical settings, which hampers their generalized use in clinical practice.

Blood-based biomarkers for Alzheimer’s disease have recently emerged as accessible, cost-effective, and relatively non-invasive tool for detecting Alzheimer’s disease neuropathology *in vivo*, aiming at circumventing the aforementioned limitations of PET and CSF biomarkers (Zetterberg, 2019). Recent studies have found that blood plasma levels of Aβ, with the Aβ42/40 ratio reflecting brain amyloid deposition, as well as levels of the neuronal injury markers neurofilament light chain (NfL) and total tau (t-tau) were significantly different in Alzheimer’s disease patients compared to healthy control individuals (Mattsson *et al*., 2016; Mattsson *et al*., 2017; Ovod *et al*., 2017; Nakamura *et al*., 2018; Schindler *et al*., 2019). They have also been shown to predict disease progression (Mielke *et al*., 2017; Ashton *et al*., 2019; Mattsson *et al*., 2019; Schindler *et al*., 2019), suggesting that these markers could potentially be used as simple and accessible tests for Alzheimer’s disease. However, NfL and t-tau, whether derived from CSF or blood, are not specific to Alzheimer’s disease (Ashton *et al*., 2020), and plasma-derived measures of Aβ yield only modest discrimination between Aβ-positive and Aβ-negative subjects as defined by validated approaches based on Aβ-PET (Nakamura *et al*., 2018; Karikari *et al*., 2020). This indicates that these blood-based markers might not be sufficiently specific or accurate to diagnose Alzheimer’s disease. In contrast, recent studies have shown that tau phosphorylated at threonine 181 (p-tau181) in plasma increases gradually across the Alzheimer’s disease continuum, accurately predicts cross-sectional brain Aβ and tau pathology as assessed with PET, and reliably discriminates Alzheimer’s disease from other neurodegenerative disorders (Mielke *et al*., 2018; Benussi *et al*., 2020; Janelidze *et al*., 2020; Karikari *et al*., 2020; Lantero-Rodriguez *et al*., 2020; Thijssen *et al*., 2020). In familial Alzheimer’s disease, the biomarker starts to increase approximately 16 years prior to estimated symptom onset (O’Connor *et al*., 2020). In direct comparisons, plasma p-tau181 was more disease-specific and accurate than the other plasma-based biomarker candidates (Janelidze *et al*., 2020; Karikari *et al*., 2020), indicating its potential as a feasible and reliable first-line test for Alzheimer’s disease in the clinic as well as in disease-modifying trials.

So far, apart from a relatively small familial Alzheimer’s disease study (O’Connor *et al*., 2020), available studies on plasma p-tau181 are limited to cross-sectional designs (Benussi *et al*., 2020; Janelidze *et al*., 2020; Karikari *et al*., 2020; Thijssen *et al*., 2020); therefore, the temporal dynamics of plasma p-tau181 changes across the spectrum of Alzheimer’s disease, as well as its associations with the temporospatial progression of Alzheimer’s disease pathology as measured by PET, remain unexplored. Addressing these questions is crucial to understand the full potential of plasma p-tau181 as an early predictor of Alzheimer’s disease, as well as to more closely elucidate the specific aspects of Alzheimer’s disease pathology reflected by this novel biomarker.

In the present study, we investigated the temporal trajectories of plasma p-tau181 across the spectrum of sporadic Alzheimer’s disease and analyzed their association with the spatiotemporal progression patterns of PET-measured Aβ and tau pathology, as well as the trajectories of established CSF biomarkers. We examined a large, prospective cohort spanning the entire clinical Alzheimer’s disease continuum with longitudinal plasma p-tau181 data as well as PET- and CSF-based biomarkers. Under the hypothesis that plasma p-tau181 is a specific marker for Alzheimer’s disease, our aims were to determine the natural course of plasma p-tau181 across the disease spectrum, investigating the specific events in the Alzheimer’s disease cascade that most closely associate with dynamic changes in plasma p-tau181, and to estimate the time point in this cascade at which plasma p-tau181 reaches abnormal levels.

## Material and methods

### Study design

Data were obtained from the Alzheimer’s Disease Neuroimaging Initiative (ADNI) database (http://adni.loni.usc.edu). The ADNI is an ongoing observational study that was launched in 2003 as a public-private partnership, led by Principal Investigator Michael W. Weiner, MD. ADNI recruits participants at 57 sites in the United States and Canada. The primary goal of ADNI has been to test whether serial magnetic resonance imaging (MRI), PET, other biological markers, and clinical and neuropsychological assessment can be combined to measure the progression of mild cognitive impairment (MCI) and early Alzheimer’s disease. The study was approved by the Institutional Review Board (IRB) of all participating centers in ADNI. All study participants, or their study partners, provided written informed consent. For the present study, data were obtained from the Laboratory of Neuro Imaging (LONI) database on June 2020.

### Participants

We included all cognitively normal (CN) participants, patients with MCI, and patients with Alzheimer’s disease dementia with at least one available plasma p-tau181 measurement and Aβ PET scan ([18F]florbetapir) at baseline (*n*=1067). A subset of these participants (*n*=864) also had available measures of Aβ_1-42_ and p-tau181 in CSF. ADNI participants were scheduled to undergo follow-up measurements of the aforementioned biomarkers (see specific Materials and Methods sections for details). Additionally, another subset of study participants (156 CN, 138 MCI, and four Alzheimer’s disease dementia) that continued in ADNI3 were scanned using tau PET imaging with [18F]flortaucipir at an average of 6.12 years after the baseline visit. Characteristics of study participants are detailed in Table 1. ADNI inclusion criteria for the diagnostic cohorts have been described in detail elsewhere (Petersen *et al*., 2010).

**Table 1.** Demographic information of study participants. Age is reported as mean (standard deviation). Continuous biomarker data are reported as median (range). MMSE was reported as median (range). Abbreviations: CN: Cognitively normal; MCI: Mild cognitive impairment; AD: Alzheimer’s disease; MMSE: Mini-mental state examination; Aβ: amyloid-β; p-tau181: phosphorylated tau at threonine 181.

|  | CN | MCI | AD |
| --- | --- | --- | --- |
| ***n*** | 359 | 518 | 186 |
| **Age, years** | 74.7 (6.7) | 72.8 (7.9) | 75.1 (7.8) |
| **Sex, M/F** | 168/191 | 291/227 | 108/78 |
| **APOE $\epsilon 4$ carriers, *n* (%+)** | 102 (28) | 241 (47) | 122 (66) |
| **MMSE** | 29 (24-30) | 28 (24-30) | 23 (9-26) |
| **Aβ positive, *n* (%)** | 110 (31) | 277 (53) | 160 (86) |
| **Centiloid (CL)** | 9.4 (-24.1 to 168.2) | 30.2 (-29.7 to 188.8) | 85.0 (-25.3 to 194.7) |
| **Plasma p-tau181, pg/ml** | 13.6 (0.8-72.3) | 15.8 (1.6-69.6) | 23.2 (6.3-63.3) |
| **CSF A$\beta_{1-42}$, pg/ml** | 1291 (203-3462) | 906 (248-3392) | 624 (255-3139) |
| **CSF p-tau181, pg/ml** | 19.6 (8.0-60.0) | 22.8 (8.2-91.3) | 33.4 (10.8-90) |

### Plasma p-tau181 measurements

Blood samples were collected and processed according to the ADNI protocol (Kang *et al*., 2015). Plasma p-tau181 concentrations were measured at the Clinical Neurochemistry Laboratory, University of Gothenburg (Mölndal, Sweden) using an assay developed in-house on a Simoa HD-X (Quanterix, Billerica, MA, USA) instrument as described previously in detail (Karikari *et al*., 2020). In brief, the AT270 mouse monoclonal antibody (MN1050; Invitrogen, Waltham, MA, USA) specific for the threonine-181 phosphorylation site, coupled to paramagnetic beads (103207; Quanterix) was used for capture and the anti-tau mouse monoclonal antibody Tau12 (806502; BioLegend, San Diego, CA, USA), which binds the N-terminal epitope 6-QEFEVMEDHAGT-18 on human tau protein for detection. All the available samples were analyzed in a single batch. We identified four participants (0.4%) with outlier values of plasma p-tau181 levels that were discarded from subsequent analyses (see Supplementary Fig. 1). Longitudinal blood sampling was performed approximately every year, over a median follow-up time of 2.9 years in 938 subjects.

### Image processing and analysis

Aβ PET imaging in ADNI was performed using [18F]florbetapir (FBP), with an injected dose of 370±37 MBq. PET images were acquired 50-70 min after injection of FBP using a dynamic protocol (4×5 minute frames). Longitudinal Aβ PET scans were acquired approximately every two years, with a median follow-up time of 4.0 years in 728 participants. Tau PET images were acquired 75-105 minutes after the injection of 370±37 MBq of [^18^F]flortaucipir (FTP) using a 6×5 minute dynamic protocol. PET preprocessing steps for scanner harmonization were identical for all the tracers and are described elsewhere (Jagust *et al*., 2015). Briefly, PET frames were realigned, averaged, reoriented, resliced to a common grid, and smoothed to a common resolution of 8 mm. Further details on PET acquisition and preprocessing in ADNI can be found at http://adni.loni.usc.edu/methods/documents/.

For quantitative PET analyses, preprocessed PET images were rigidly co-registered to the closest-in-time corresponding structural T1 MRI scan using Statistical Parametric Mapping 12 (SPM12; Wellcome Department of Imaging Neuroscience, Institute of Neurology, London, UK). T1 MRI acquisition protocols and standardized preprocessing steps for scanner harmonization and noise reduction have been described earlier (Jack *et al*., 2015). The preprocessed T1 MRI scan was then automatically segmented into grey (GM) and white matter (WM) tissue segments and high-dimensionally registered to Montreal Neurological Institute (MNI) space using the Computational Anatomy Toolbox (CAT12, http://dbm.neuro.uni-jena.de/cat/) in SPM12. Binary GM and WM masks were created using a threshold of 0.5 over the corresponding tissue probability map in participant space. The inverse of the deformation field resulting from spatial registration was used to propagate regions of interest (ROI) from MNI to participant space, and the propagated ROIs were multiplied by the appropriate binary segment to create the final mask. We generated standardized uptake value ratio (SUVR) images for FBP using a whole cerebellum ROI (Klunk *et al*., 2015) as the reference region. Global Aβ deposition was defined as the mean SUVR in a previously defined cortical composite ROI (Klunk *et al*., 2015), and these values were then transformed to Centiloid units (Klunk *et al*., 2015) using equations derived by the ADNI PET Core (http://adni.loni.usc.edu/data-samples/access-data/). FBP SUVR images were finally corrected for partial volume effects (PVE) using the Müller-Gärtner method (Gonzalez-Escamilla *et al*., 2017). For FTP imaging, SUVR maps were created using an inferior cerebellum ROI (Maass *et al*., 2017) as the reference region and corrected for PVE using the region-based voxel-wise (RBV) method (Thomas *et al*., 2011) with a previously defined anatomical parcellation (Baker *et al*., 2017). To perform voxel-wise analyses, co-registered PET images were spatially normalized to MNI space using the deformation field obtained from spatial normalization of their corresponding MRI scan, and the resulting images were masked with a GM mask and smoothed using a 6 mm isotropic filter.

### CSF biomarkers

CSF samples were collected and processed according to previously described protocols (Kang *et al*., 2015). Concentrations of Aβ_1-42_ and p-tau181 in CSF were measured by the ADNI Biomarker Core using the Elecsys® β-Amyloid(1–42) and the Elecsys® Phospho-Tau (181P) CSF immunoassays, respectively, on a cobas e 601 module (Bittner *et al*., 2016; Hansson *et al*., 2018). The measuring limits (lower to upper limits) of these assays are 200 to 1700 ng/l for Elecsys® β-Amyloid(1–42) and 8 to 120 ng/l for Elecsys® Phospho-Tau (181P) assays. The measuring range of the Elecsys® β-Amyloid(1-42) CSF immunoassay beyond the upper technical limit has not been formally established. Therefore, use of values above the upper technical limit, which are provided based on an extrapolation of the calibration curve, is restricted to exploratory research purposes and is excluded for clinical decision making or for the derivation of medical decision points. In the present study, we included extrapolated values of Aβ_1-42_ concentrations in all the analyses. Longitudinal CSF extractions were performed approximately every two years over a median follow-up time of 3.3 years in 410 participants.

### Biomarker cut-points

To determine Aβ status (+/-) using Aβ PET, we used an externally derived cut-point of 24.4 Centiloids that best discriminated between subjects with and without Alzheimer’s disease neuropathologic changes at autopsy (La Joie *et al*., 2019). The cut-point for CSF Aβ_1-42_ +/- using the Elecsys assay was also independently determined on the basis of maximal agreement with Aβ PET (1100 pg/ml) (Hansson *et al*., 2018; Schindler *et al*., 2019). No externally determined cut-points for p-tau181 markers (CSF and plasma) are currently available, and therefore we derived these cut-points conservatively by defining them as the 90^th^ percentile of PET-Aβ-CN individuals (*n*=190) (Jack *et al*., 2017), yielding 21.99 pg/ml for plasma p-tau181 and 28.69 for CSF p-tau181.

### Statistical analysis

Longitudinal rates of change in plasma p-tau181 levels as well as PET-derived Centiloid values and CSF biomarker levels were estimated using linear mixed models with subject-specific intercepts and slopes that predicted biomarker levels over time (*biomarker∼time+(time*|*subject)*). Individual rates of change were derived from these models by summing the fixed and the subject-specific random effects terms and were used for subsequent analyses as described below.

We first assessed linear associations of baseline plasma p-tau181 levels with cross-sectional and longitudinal estimates of regional Aβ accumulation as measured by FBP-PET, using voxel-wise linear regressions adjusted for age and sex. Identical models were used to assess associations between longitudinal changes in plasma p-tau181 and increases in voxel-wise FBP-PET signal. In order to assess a possible disease stage dependency of these associations all models were computed for the different diagnostic groups separately.

Second, we used non-linear smoothing spline regressions to model baseline levels and longitudinal changes in plasma p-tau181 as a function of globally increasing Aβ pathology, both measured using CSF Aβ_1-42_ levels and FBP-PET-derived Centiloid values. The smoothing parameter was determined via minimization of the mean squared error using a 25-repetition, 10-fold cross-validation procedure. 95% confidence intervals (CI) were generated using 5000-repetition bootstrap samples. This procedure was also employed to describe the dependency between baseline levels and longitudinal change of p-tau181 as measured in plasma and CSF.

Third, we assessed associations of baseline levels and longitudinal changes in plasma p-tau181 with future tau deposition measured on FTP PET six years later, using linear voxel-wise regressions adjusted for age, sex, and time difference between FTP scan and blood extraction. Analyses were conducted separately for cognitively normal and cognitively impaired individuals (pooled MCI + Alzheimer’s disease dementia due to the low number of Alzheimer’s disease dementia patients).

Finally, we aimed at determining the temporal trajectories of plasma p-tau181 and core Alzheimer’s disease biomarkers across the spectrum of sporadic Alzheimer’s disease. For this purpose, we employed a previously developed method for the construction of long-term temporal biomarker trajectories using individual short-term data (Villemagne *et al*., 2013; Budgeon *et al*., 2017). Briefly, annualized rates of change were plotted against their corresponding baseline levels, transformed to z-scores with reference to Aβ-CN subjects, and fitted using the above described smoothing spline procedure. Resulting spline curves were finally integrated using Euler’s method and anchored to z-score=0 at t=0, therefore describing the temporal evolution of biomarkers from characteristic levels of subjects without evident Aβ pathology to fully abnormal levels.

### Data availability

Data used in this study has been made publicly available by the ADNI in the Laboratory of Neuro Imaging (LONI) database.

## Results

### Associations of plasma p-tau181 with regional Aβ pathology across the Alzheimer’s clinical spectrum

First, we assessed the cross-sectional associations of plasma p-tau181 with global and regional Aβ deposition on FBP-PET across the clinical spectrum of Alzheimer’s disease (Fig. 1A). Baseline levels of plasma p-tau181 associated with Aβ deposition more strongly in subjects with MCI and Alzheimer’s disease dementia, while associations were markedly weaker among CN participants. The observed association patterns in MCI and Alzheimer’s disease dementia subjects covered widespread areas of the cortex and expanded sub-cortically to the striatum (see Supplementary Fig. 2). In contrast, the weaker associations observed in CN subjects were restricted to the precuneus and to temporal and superior-frontal areas, and did not involve subcortical structures, suggesting that plasma p-tau181 associates more strongly with Aβ pathology when Aβ deposits are present in widespread areas of the brain. Results were similar when using global composite PET-imaging measures of Aβ pathology (Fig. 1A, right panel, *r*=0.18 in CN, *r*=0.41 in MCI, and *r*=0.35 in Alzheimer’s disease, *p*<0.001 for all age- and sex-adjusted associations). We then investigated the correlations of baseline and change measures of plasma p-tau181 with longitudinal Aβ accumulation in serial FBP-PET (Figs. 1B and 1C). Strongest associations were again observed in MCI participants, followed by CN subjects. Only marginal and statistically non-significant associations were found for Alzheimer’s disease dementia patients, which, however, also had a much smaller sample size. Similar to the cross-sectional findings, regional association patterns in CN and MCI individuals revealed that both elevated baseline levels and longitudinal increases of plasma p-tau181 were associated with longitudinal Aβ accumulation in large areas of the temporal, frontal, and parietal cortices, as well as in the striatum (see Supplementary Fig. 3), which suggests a stronger association of plasma p-tau181 with Aβ in advanced stages of brain amyloidosis. Associations with global longitudinal Aβ accumulation were statistically significant for CN (*r*=0.17, *p*=0.006 for baseline plasma p-tau181, *r*=0.22, *p*<0.001 for change in plasma p-tau181) and MCI (*r*=0.27 for baseline, *r*=0.26 for change, *p*<0.001 for both) but not for Alzheimer’s disease dementia subjects (*r*=0.23, *p*=0.12 for baseline and *r*=0.03, *p*=0.79 for change).

**Figure 1.**
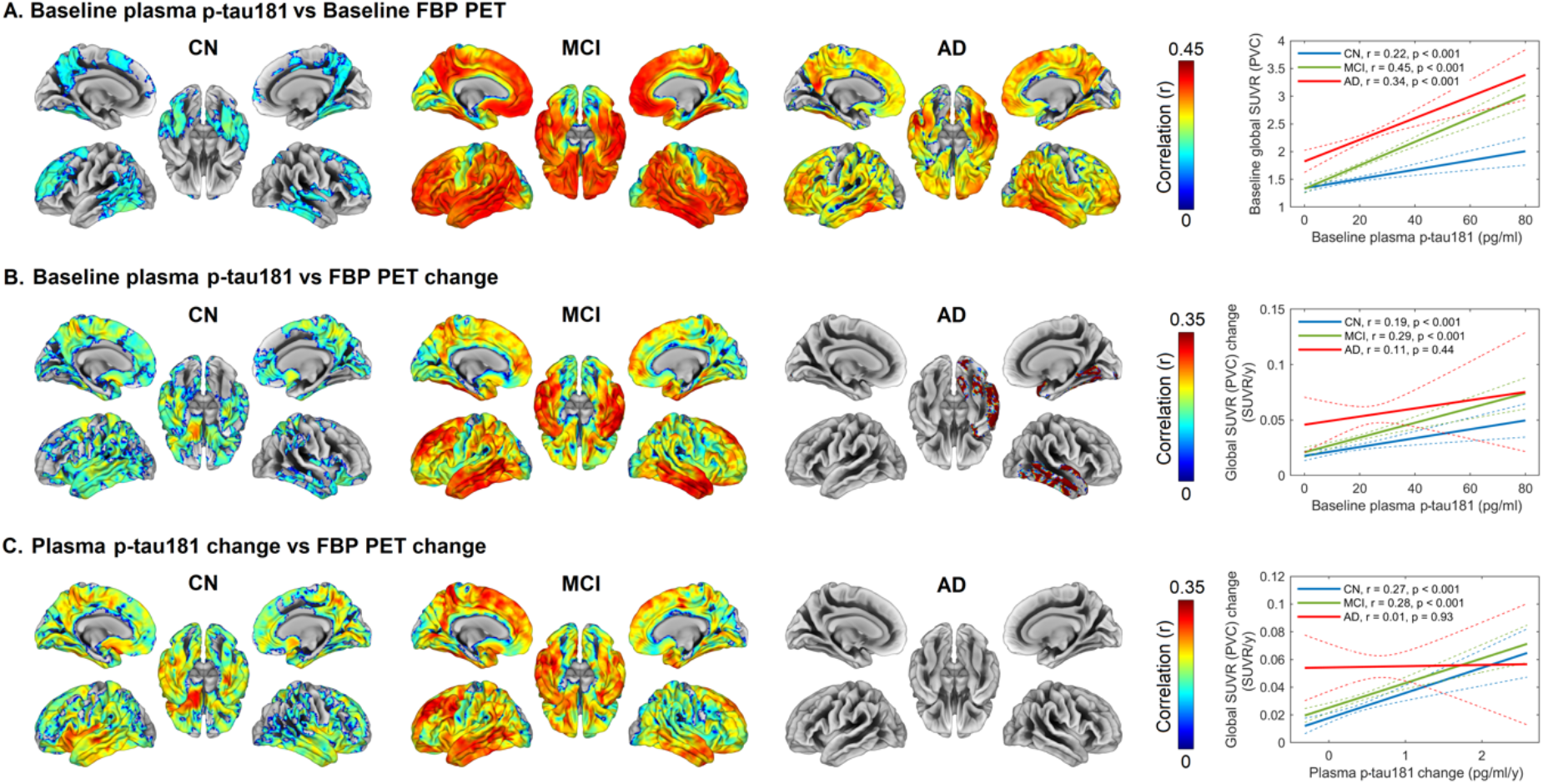
Regional and global associations of plasma p-tau181 with PET-measured Aβ deposition and longitudinal accumulation across the clinical spectrum of Alzheimer’s disease. Voxel-wise analyses (adjusted for age and sex) assessing regional associations between (A) baseline plasma p-tau181 levels and baseline FBP SUVR, (B) baseline plasma p-tau181 levels and FBP SUVR change, and (C) plasma p-tau181 change and FBP SUVR change. Significant associations in voxel-wise analyses were determined based on voxel-level thresholds of *p*_uncorrected_<0.001 (A), or *p*_uncorrected_<0.01 (B) and (C), and a family-wise error (FWE)-corrected threshold of *p*<0.001 at the cluster level for all analyses. Color panels on the right display linear fits and (unadjusted) Pearson correlation coefficients (r) of the effects on global measures. Dashed lines in right panels are 95% confidence intervals. In CN, weak correlations were observed in regions previously described as early Aβ accumulating regions, while the strongest correlations were observed at the MCI stage where regional associations covered widespread areas involving cortical and subcortical areas known to be involved later in the disease course (see also Supplementary Fig. 2 and Supplementary Fig. 3). AD: Alzheimer’s disease.

### Plasma p-tau181 dynamic changes and Aβ pathology

Using spline regression, we observed that earliest elevations in baseline plasma p-tau181 levels as a function of global Aβ pathology occurred even before FBP PET and CSF Aβ_1-42_ reached their respective abnormality thresholds (Fig. 2A and Fig. 2B, left panels), demonstrating consistent increases as Aβ pathology progresses. This cross-sectional result was confirmed when analysing the dependence of plasma p-tau181 change rates on Aβ biomarker levels: a small but significant deviation from normative levels in the standardized change rate was observed at subthreshold levels for both FBP-PET and CSF Aβ_1-42_ (Fig. 2A and Fig. 2B, right panels) and this change continued accelerating as the severity of Aβ pathology increased. The cut-point for abnormal levels of plasma p-tau181, as defined above, was reached at relatively advanced levels of global Aβ pathology (Centiloid=70 and CSF Aβ_1-42_=540 pg/ml), confirming our previous regional neuroimaging analyses.

**Figure 2.**
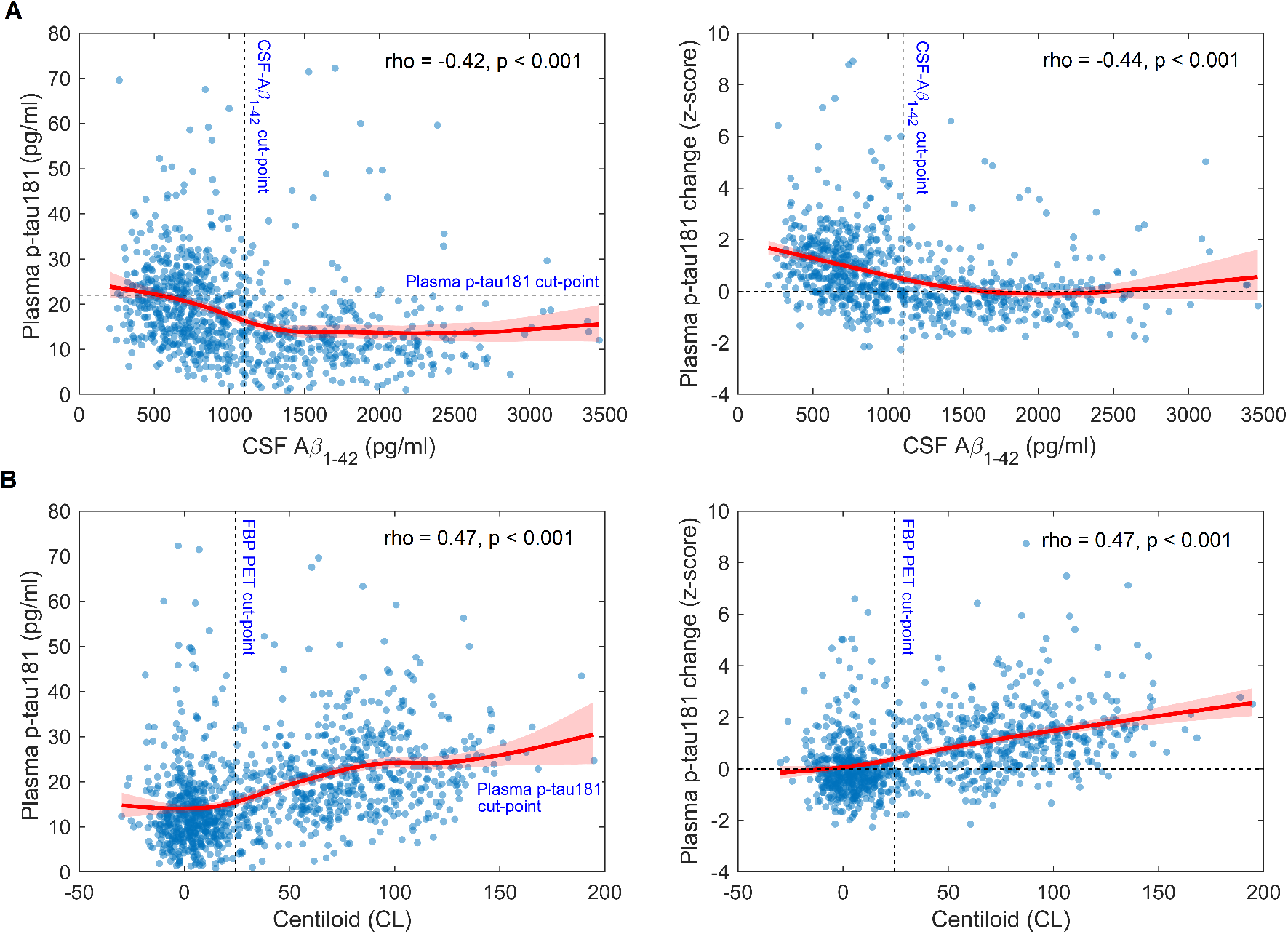
Baseline and longitudinal associations of plasma p-tau181 with imaging and CSF biomarkers of global Aβ pathology. Spline regressions describing the statistical dependence of baseline levels of plasma p-tau181 (left-side panels) and longitudinal change in plasma p-tau181 (right side panels) on (A) CSF Aβ1-42 levels and (B) global FBP SUVR. Spearman’s rank correlation was used to quantify the monotonic correlation between these measures. Shaded areas are 95% confidence intervals for the fit. Dashed lines represent cut-points for abnormality for the studied biomarkers. Earliest increases in plasma p-tau181 appeared shortly before Aβ markers reached abnormal levels, and changes accelerated as the severity of global Aβ pathology increased. Plasma p-tau181 reached abnormal levels only after Aβ biomarkers reached abnormal levels (PET Centiloid=70 and CSF Aβ1-42=540 pg/ml, see left-side panels).

### Associations between p-tau181 levels in plasma and in CSF

Spline regressions demonstrated that cross-sectional p-tau181 levels in plasma and in CSF were strongly correlated up to relatively high levels of CSF p-tau181 (∼50 pg/ml) (Fig. 3A). Moreover, longitudinal increases in plasma p-tau181 were found to accelerate with increasing baseline CSF p-tau181 levels (Fig. 3B). Finally, the p-tau181 change rates in plasma and in CSF followed a linear trend approximately anchored at z-scores (0,0), indicating that these two measures follow similar dynamics (Fig. 3C).

**Figure 3.**
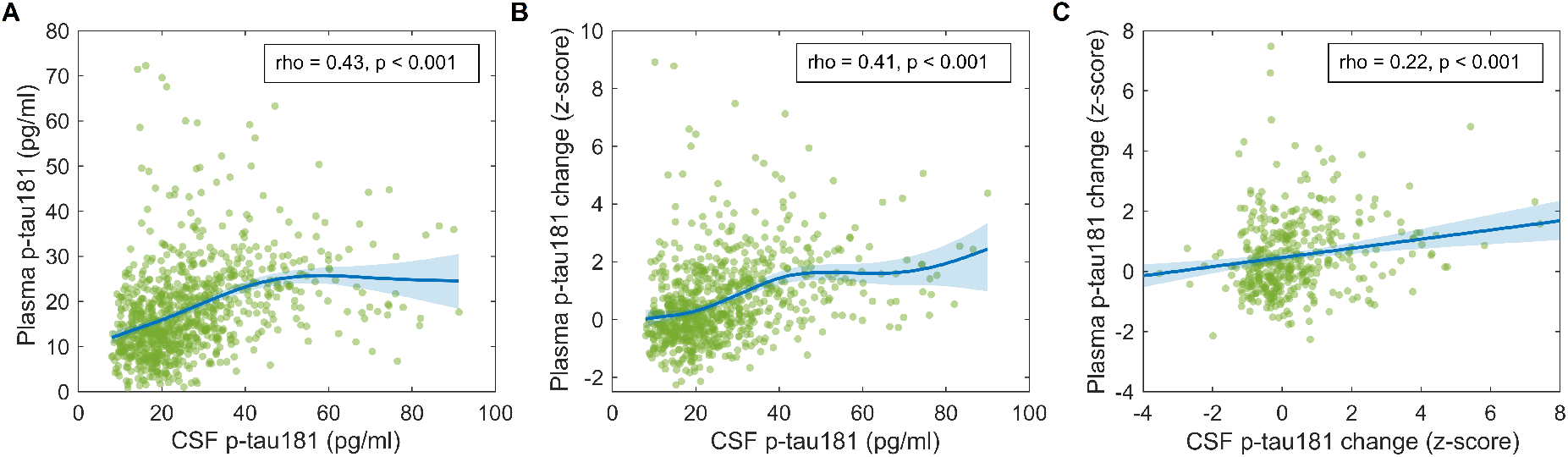
Baseline and longitudinal associations between plasma p-tau181 and CSF p-tau181. Spline regressions describing the statistical dependence of baseline levels of plasma p-tau181 (A) and plasma p-tau181 change (B) on CSF p-tau181 levels, as well as plasma p-tau181 change vs CSF p-tau181 change (C). Z-scores were computed using Aβ-CN levels as the reference. Shaded areas are 95% confidence intervals for the fit. Spearman’s rank correlation was used to quantify the monotonic correlation between these measures. Changes of p-tau181 in plasma and CSF followed a linear trend approximately anchored at the origin, indicating that these two markers follow similar dynamics.

### Associations between plasma p-tau181 and regional tau deposition six years later

We then investigated whether baseline and change measures of plasma p-tau181 correlated with the severity of PET-measured tau pathology six years later (Fig. 4). In CN subjects, baseline plasma p-tau181 correlated with future tau pathology in brain regions mainly restricted to the medial temporal and posterior cingulate cortex (Fig. 4A). In cognitively impaired individuals, associations were stronger and statistically significant in broader areas of the cortex, particularly in lateral temporo-parietal cortical areas. Compared to baseline measures, longitudinal increase in plasma p-tau181 was even stronger associated with brain tau pathology six years later, particularly in CN individuals (Fig 4b). Thus, significant associations in both CN and cognitively impaired individuals were observed across a pronounced temporo-parietal cortical pattern that closely resembled the stereotypical spatial pattern of NFT aggregation in Alzheimer’s disease.

**Figure 4.**
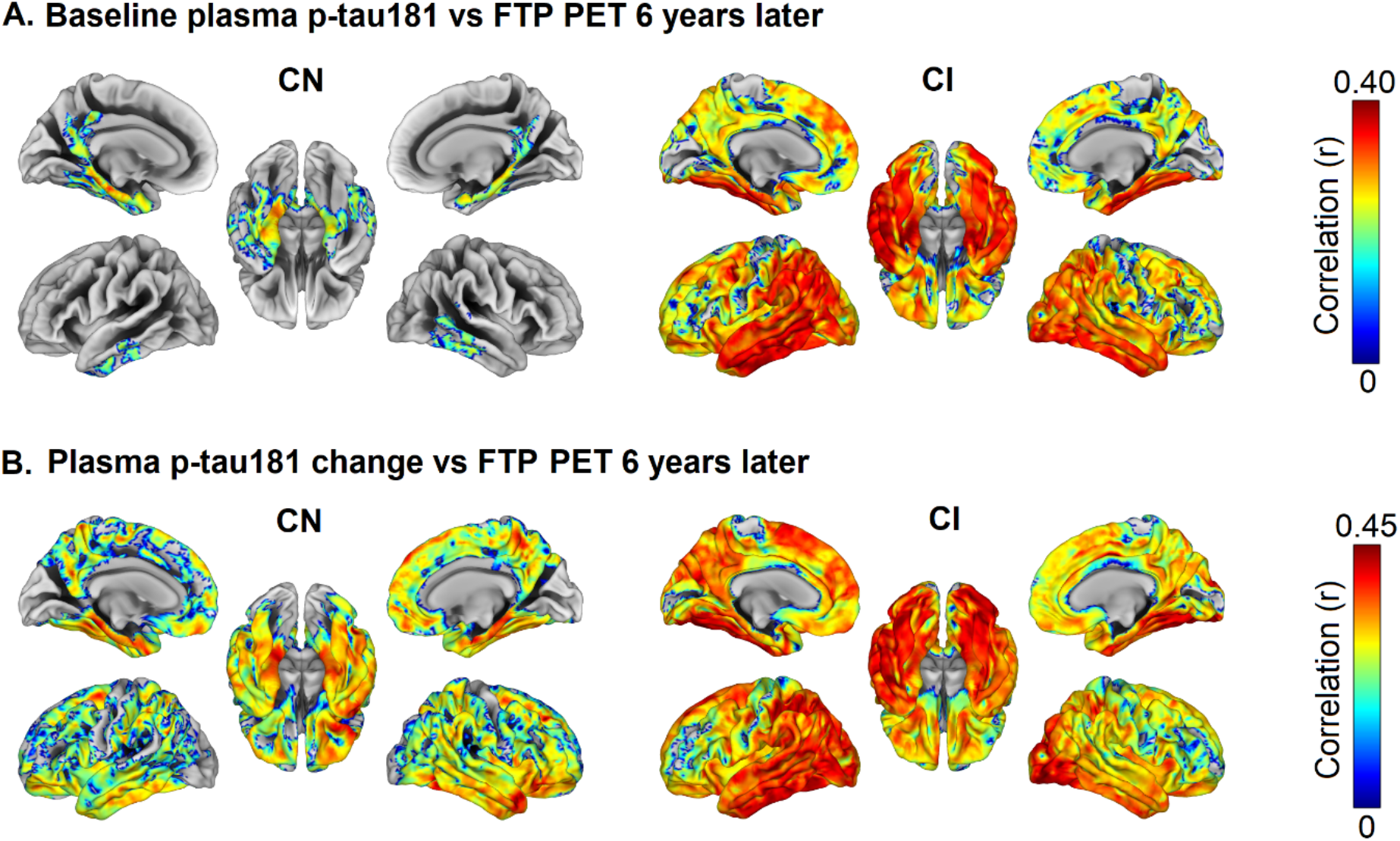
Associations of plasma p-tau181 baseline levels and longitudinal increase with regional tau aggregation six years later. Regional associations (adjusted for age, sex, and time difference between blood sampling and PET scanning) of (A) baseline plasma p-tau181 levels and (B) longitudinal plasma p-tau181 change with voxel-wise FTP SUVR six years later. Significant associations in voxel-wise analyses were determined based on a voxel level threshold of *p*_uncorrected_<0.01 and a family-wise error (FWE)-corrected threshold of *p*<0.001 at the cluster level. Baseline plasma p-tau181 and, more pronounced, longitudinal change were associated with widespread tau aggregation six years later, following the characteristic temporo-parietal pattern of progressing neurofibrillary tangle pathology in Alzheimer’s disease. CI: cognitively impaired.

### Temporal trajectories of plasma p-tau181 in comparison to established Alzheimer’s disease biomarkers

We finally determined the temporal trajectory followed by plasma p-tau181 levels and compared it to the trajectories of established PET and CSF Alzheimer’s disease biomarkers. Spline regressions demonstrated that plasma p-tau181 and both PET- and CSF-based Aβ biomarkers rates of change showed an inverted-U shaped dependence on baseline values (Fig. 5A), indicating that change rates for these markers decelerate for highly abnormal baseline levels. In contrast, CSF p-tau181 change increased in a linear manner over the entire range of baseline values. Spline regressions in Fig. 5A were integrated and anchored at median levels of Aβ-CN to derive comparative temporal trajectories of plasma p-tau181, FBP-PET, and CSF biomarkers (Fig 5B). Plasma p-tau181 reached abnormal levels after 23.2 (95% CI: 22.0 to 22.4) years (Fig 5b, left panel), significantly later than CSF Aβ_1-42_ (16.7 years, difference -6.5, 95% CI: -8.6 to -4.1) and FBP-PET (17.5 years, difference -5.7 years, 95% CI: -7.8 to -3.2) (Fig. 5B, right panel). Plasma p-tau181 and CSF p-tau181 followed very similar trajectories, the latter reaching abnormal levels 2 years after plasma p-tau181, although this difference was not statistically significant (95% CI: -0.2 to 4.65).

**Figure 5.**
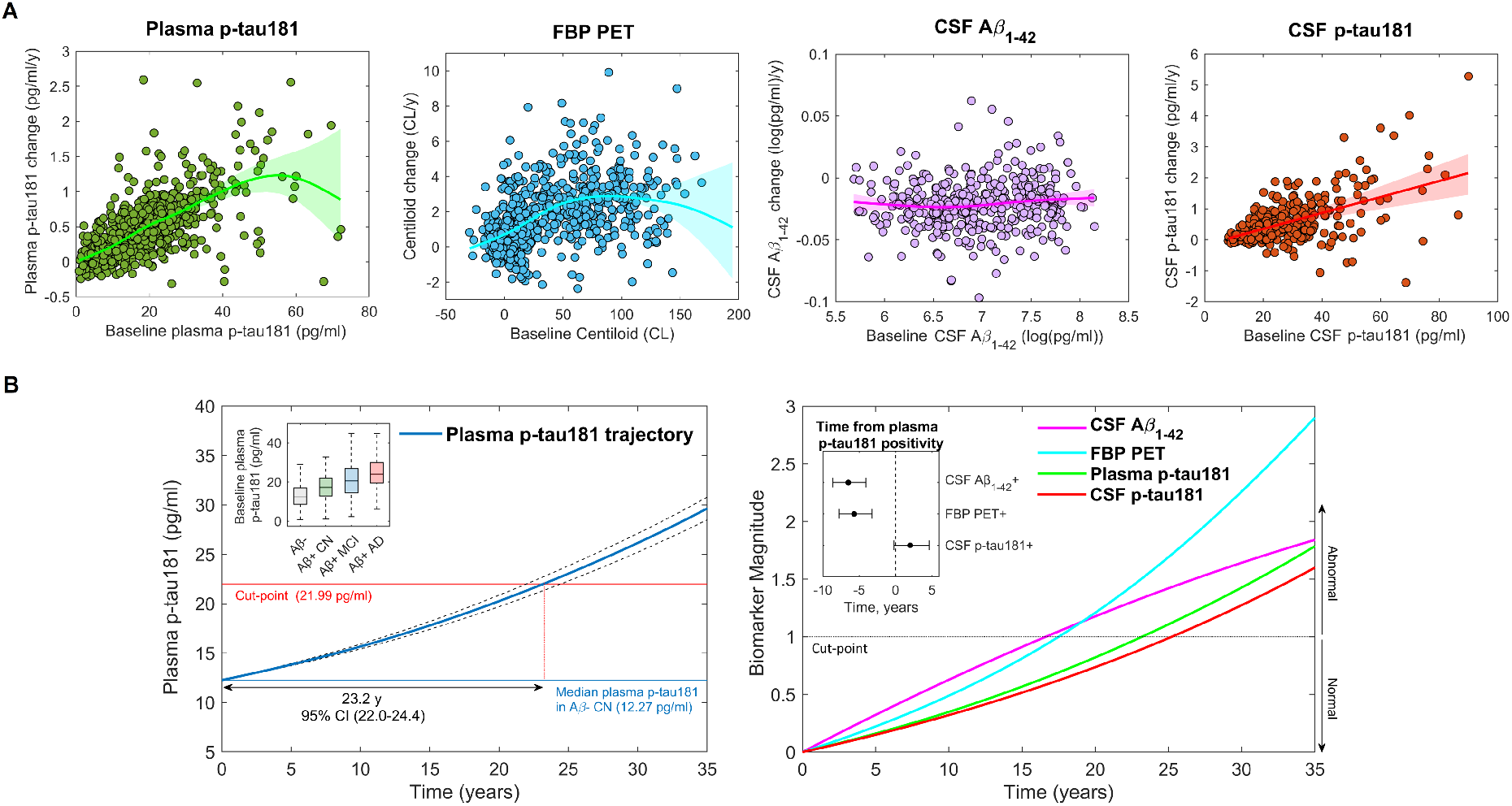
The natural time course of plasma p-tau181 in Alzheimer’s disease. (A) Spline regressions describing the dependence of biomarker rate of change on baseline levels of the respective biomarker. Logarithmic transformation of CSF Aβ1-42 levels was performed in order to improve residual normality in linear mixed models and facilitate the spline fit. (B) Left panel: Time course of plasma p-tau181, estimated using individual longitudinal data. The curve is anchored at median plasma p-tau181 levels in CN Aβ-individuals, thus describing the temporal trajectory from non-pathologic to abnormal levels. The inner panel shows a box plot representing biomarker levels for Aβ- and Aβ+ subjects at different stages of Alzheimer’s disease. Right panel: Combined temporal trajectories of plasma p-tau181, Aβ PET and CSF biomarkers. To represent all trajectories on the same scale, curves were anchored to median levels in CN Aβ-subjects, transformed to z-scores using mean CN Aβ-levels as reference, and scaled to the corresponding cut-point z-score (Villemagne et al., 2013). The inner panel demonstrates the time lag between time points where plasma p-tau181 and other biomarkers reach abnormal levels. Plasma p-tau181 reached abnormal levels approximately 5.7 years after Aβ PET and 6.5 years after CSF Aβ1-42, following similar dynamics as CSF p-tau181, which reached abnormal levels 2.0 years after plasma p-tau181.

## Discussion

In the present prospective longitudinal study, we provide a detailed description of the temporal dynamics of plasma p-tau181, from the earliest manifestations of Alzheimer’s disease pathology in cognitively normal individuals to the dementia stage. Our findings indicate that 1) established Aβ pathology associates with dynamic changes of p-tau181 in blood, 2) elevated levels of plasma p-tau181 associate with elevated tau-PET signal six years later, with a spatial distribution that closely matches the typical predilection sites of NFT pathology in Alzheimer’s disease, and 3) p-tau181 in blood largely reflects the dynamics of p-tau181 in CSF. Taken together, these results suggest that plasma p-tau181 reflects features of tau pathology that are intimately related to fibrillar Aβ pathology and that might be predictive of downstream aggregation of tau fibrils several years before established NFT pathology. Our findings thus extend prior results from three recent cross-sectional studies (Janelidze *et al*., 2020; Karikari *et al*., 2020; Thijssen *et al*., 2020) and from a study in familial Alzheimer’s disease (O’Connor *et al*., 2020), providing a more comprehensive picture of the pathological processes reflected by this ultrasensitive measure of p-tau181 in blood. Moreover, by analyzing longitudinal changes on multimodal biomarker data, we determined, for the first time, the precise sequence of pathological events that accompany the natural course of plasma p-tau181 changes across the spectrum of Alzheimer’s disease.

First studies of the novel plasma p-tau181 assays could demonstrate a good correspondence of plasma p-tau181 levels with a positive Aβ and tau status as measured with PET (Janelidze *et al*., 2020; Karikari *et al*., 2020; Thijssen *et al*., 2020). However, the strength of the associations with PET-measured Aβ and tau pathologies was not consistent across studies, with two studies showing stronger associations with Aβ (Karikari *et al*., 2020; Thijssen *et al*., 2020) and the third with tau (Janelidze *et al*., 2020), leaving unclear the specific pathologic process best reflected by plasma p-tau181. Moreover, these studies were limited by their cross-sectional design and could not provide direct insights into the temporal dynamics of plasma p-tau181 changes in relation to established PET- and CSF-based biomarkers of Alzheimer’s disease pathology. Therefore, the ability of plasma p-tau181 to detect early pathologic features of the disease was not clear.

A first key finding of our longitudinal study describes that the earliest dynamic changes in plasma p-tau181 levels occurred even before PET and CSF biomarkers for Aβ pathology reached abnormal levels (Fig. 2). Moreover, the dynamics of plasma p-tau181 increases accelerated as the severity of Aβ pathology increased, reaching abnormal levels approximately 5 years after established Aβ pathology (Fig. 5). Consistent with these findings, we found that the associations of plasma p-tau181 with regional Aβ deposition as measured with FBP-PET were stronger when Aβ deposits had significantly spread throughout the cortex (Fig. 1). Further, plasma p-tau181 changes were associated with longitudinal Aβ accumulation in several cortical and subcortical areas that have been previously identified as late-accumulating areas in the disease course (Thal *et al*., 2002; Grothe *et al*., 2017; Palmqvist *et al*., 2017; Hanseeuw *et al*., 2018). Plasma p-tau181 associations with Aβ accumulation were marginal among Alzheimer’s disease dementia patients, suggesting Aβ saturation effects at this stage (Jack *et al*., 2013). Overall, these results suggest that early Aβ pathology associates with tau dysregulation and subsequent release of soluble p-tau181 in blood, which escalates at more advanced Aβ stages. This is in line with previous *in-vitro* (De Felice *et al*., 2008; Jin *et al*., 2011) and *in-vivo* animal findings (Zheng *et al*., 2002; Shin *et al*., 2007), as well as results from a recent report on p-tau181 in CSF (Mattsson-Carlgren *et al*., 2020).

A second key finding of our study revealed that baseline plasma p-tau181 levels and, more pronounced, longitudinal plasma p-tau181 increases, were associated with PET-measured tau aggregation six years later (Fig. 4), suggesting that the progressive accumulation of soluble p-tau181 might be a marker of tau fibril aggregation. Interestingly, baseline plasma p-tau181 levels even predicted spatially restricted tau aggregation in cognitively normal individuals, coinciding with typical limbic predilection sites of initial NFT formation (Braak *et al*., 2006; Brier *et al*., 2016; Johnson *et al*., 2016; Schöll *et al*., 2016; Bejanin *et al*., 2017; Hanseeuw *et al*., 2019). By contrast, dynamic increases in plasma p-tau181 levels correlated with NFT pathology in widespread cortical areas exceeding the medial temporal lobe, following a typical temporo-parietal distribution pattern characteristic for NFT deposition associated with advanced Braak stages (Braak *et al*., 2006; Schöll *et al*., 2019). These findings extend results from previous studies showing cross-sectional associations between elevations in plasma p-tau181 and widespread PET-measured NFT deposition (Janelidze *et al*., 2020; Karikari *et al*., 2020; Thijssen *et al*., 2020), demonstrating the potential of plasma p-tau181 as an accessible measure of pathological features of Alzheimer’s disease that relate more closely with clinical decline (Brier *et al*., 2016; Bejanin *et al*., 2017; Hanseeuw *et al*., 2019; Janelidze *et al*., 2020; Karikari *et al*., 2020).

In line with recent cross-sectional studies, we found strong associations between p-tau181 levels in blood and CSF (Janelidze *et al*., 2020; Karikari *et al*., 2020). Further, we extended these previous observations by noting that p-tau181 in blood and CSF followed similar longitudinal dynamics (Figs. 3C and 5B), suggesting that elevations of plasma p-tau181 in blood and CSF reflect comparable underlying pathological processes. Still, differences in diagnostic performance and predictive power between these two p-tau181 markers remain to be elucidated and are currently the focus of an ongoing investigation by our group.

Our findings have clear implications for the use of plasma p-tau181 as a diagnostic test for early Alzheimer’s disease. First, the observation that prominent changes in plasma p-tau181 coincide with the presence of established Aβ pathology indicates that these elevations are highly specific for Alzheimer’s disease neuropathologic changes. Second, plasma p-tau181 levels reached abnormality thresholds approximately five years after manifest brain amyloidosis as detected by Aβ PET or CSF Aβ_1-42_. Since overt neurodegeneration and cognitive decline occur many years (even decades) after Aβ positivity is reached (Villemagne *et al*., 2013; Baek *et al*., 2020), this indicates that, from a clinical perspective, plasma p-tau181 can be regarded as an early biomarker for Alzheimer’s disease. Third, the ability of plasma p-tau181 and its longitudinal accumulation to forecast widespread tau tangle deposition suggests that this marker might be suitable to track Alzheimer’s disease progression up to advanced disease stages that strongly associate with cognitive decline.

The results presented in this study may also have relevant implications for disease-modifying treatment trials of Alzheimer’s disease. One of the main potential applications of plasma p-tau181 is its use as a screening tool prior to Aβ or tau PET confirmatory scans, likely resulting in highly reduced costs (Jack, 2020). In this regard, our findings indicate that, although coinciding with an early clinical stage, elevated plasma p-tau181 levels associate to a disease stage of several years of pathologic disease progression that likely reflects an established disruption of tau metabolism leading to NFT formation. Thus, patient selection based on plasma p-tau181 might be detrimental for trials that target earlier features of the disease such as early Aβ pathology (Sperling *et al*., 2014). Nevertheless, the fact that plasma p-tau181 correlated with the severity of tau pathology several years later suggests that this marker might be particularly useful for screening participants in clinical trials targeting tau pathology (Congdon and Sigurdsson, 2018). Future studies are warranted to elucidate the power of plasma p-tau181 as an estimator of target engagement in tau trials.

Strengths of our study include using data from a large, prospective cohort with multimodal biomarker data to explore the associations between plasma p-tau181 and established biomarkers of different aspects of Alzheimer’s disease pathology in a relatively unbiased manner. Second, we used a longitudinal design with comparably comprehensive and long follow-up data that allowed the derivation of a robust estimate of the temporal trajectory of plasma p-tau181 changes in direct comparison to those of established Alzheimer’s disease biomarkers. Limitations include 1) tau PET was not acquired concurrently to plasma p-tau181 and therefore we could only assess associations between plasma p-tau181 and regional tau deposition six years later, whereas the baseline levels of tau deposition remain unknown. Effects of plasma p-tau181 changes on regional tau accumulation rates will have to be studied in more detail using serial PET data; 2) the estimated time point at which plasma p-tau181 levels reach abnormality in the temporal trajectory models (approx. 5y from Aβ positivity) obviously depends on the employed cut-off for denoting abnormality. Since no universally accepted cut-points for plasma or CSF p-tau181 levels are currently available in the literature we used a commonly used method for cutoff derivation in the biomarker field based on the distribution in a non-pathological (in this case Aβ-) control population (Jack *et al*., 2017). While more research on optimal plasma p-tau181 cut-offs is necessary, we note that this method yielded a cut-off for CSF p-tau181 that was very similar to previously proposed cut-offs for this biomarker (Blennow *et al*., 2019; Mattsson *et al*., 2019; Meyer *et al*., 2020). Moreover, the conservative nature of our approach ensures maximal specificity, which is the most desirable feature of this biomarker from a clinical perspective; 3) high dropout rates in the Alzheimer’s disease dementia group limited our statistical power to detect regional associations with longitudinal Aβ pathology. However, several previous studies have indicated little dynamic Aβ changes at this disease stage (Villemagne *et al*., 2011; Jack *et al*., 2013; Villemagne *et al*., 2013); 4) only four subjects with Alzheimer’s disease dementia were scanned using tau PET, leaving unclear how plasma p-tau181 specifically associates with tau pathology in this advanced disease stage; 5) the ADNI is a highly preselected cohort, which, for example, did not include participants with significant vascular pathologies. Our findings can thus not easily be extrapolated to the population at large, and possible effects of vascular pathology and other common comorbidities on plasma p-tau181 levels remain to be studied in less selected cohorts.

In conclusion, we provide a detailed picture of the temporal trajectory followed by plasma p-tau181 in the context of established Alzheimer’s disease biomarkers, in which elevations of plasma p-tau181 are tightly linked to established Aβ pathology. Moreover, dynamic changes of plasma p-tau181 closely resembled those of CSF p-tau181, suggesting that both markers reflect similar underlying pathological processes. Finally, plasma p-tau181 levels associated with advanced regional tau deposition as detected by PET several years after the blood test. Together, these findings strongly support the use of this novel blood biomarker as a diagnostic and screening tool for Alzheimer’s disease.

## Data Availability

Data used in preparation of this article were obtained from the Alzheimer's Disease Neuroimaging Initiative (ADNI) database (adni.loni.usc.edu). As such, the investigators within the ADNI contributed to the design and implementation of ADNI and/or provided data but did not participate in analysis or writing of this report. Data used in this study have been made publicly available by the ADNI in the Laboratory of Neuro Imaging (LONI) database.

## Abbreviations

Aβ: Amyloid-β
ADNI: Alzheimer’s Disease Neuroimaging Initiative
CAT12: Computational Anatomy Toolbox
CI: Confidence interval
CN: Cognitively normal
CSF: Cerebrospinal fluid
FBP: [18F]florbetapir
FTP: [18F]flortaucipir
GM: Grey matter
MCI: Mild cognitive impairment
MNI: Montreal neurological institute
MRI: Magnetic resonance imaging
NfL: Neurofilament light chain
NFT: Neurofibrillary tangles
PET: Positron emission tomography
PVE: Partial volume effects
p-tau181: tau phosphorylated at threonine 181
ROI: Region of interest
SUVR: Standardized uptake value ratio
t-tau: total tau
WM: White matter

## Acknowledgments / Funding

MJG is supported by the “Miguel Servet” program [CP19/00031] of the Spanish Instituto de Salud Carlos III (ISCIII-FEDER). TKK holds a research fellowship from the Brightfocus Foundation (#A2020812F), and is further supported by the Swedish Alzheimer Foundation (Alzheimerfonden; #AF-930627), the Swedish Brain Foundation (Hjärnfonden; #FO2020-0240), the Swedish Dementia Foundation (Demensförbundet), the Agneta Prytz-Folkes & Gösta Folkes Foundation (#2020-00124), the Aina (Ann) Wallströms and Mary-Ann Sjöbloms Foundation, the Anna Lisa and Brother Björnsson’s Foundation, Gamla Tjänarinnor, and the Gun and Bertil Stohnes Foundation. AS is supported by the Paulo Foundation and the Orion Research Foundation. MSC received funding from the European Union’s Horizon 2020 Research and Innovation Program under the Marie Sklodowska-Curie action grant agreement No 752310, and currently receives funding from Instituto de Salud Carlos III (PI19/00155) and from the Spanish Ministry of Science, Innovation and Universities (Juan de la Cierva Programme grant IJC2018-037478-I). HZ is a Wallenberg Scholar supported by grants from the Swedish Research Council (#2018-02532), the European Research Council (#681712), Swedish State Support for Clinical Research (#ALFGBG-720931), the Alzheimer Drug Discovery Foundation (ADDF), USA (#201809-2016862), and the UK Dementia Research Institute at UCL. HZ has served at scientific advisory boards for Denali, Roche Diagnostics, Wave, Samumed, Siemens Healthineers, Pinteon Therapeutics and CogRx, has given lectures in symposia sponsored by Fujirebio, Alzecure and Biogen, and is a co-founder of Brain Biomarker Solutions in Gothenburg AB (BBS), which is a part of the GU Ventures Incubator Program (outside submitted work). KB is supported by the Swedish Research Council (#2017-00915), the Alzheimer Drug Discovery Foundation (ADDF), USA (#RDAPB-201809-2016615), the Swedish Alzheimer Foundation (#AF-742881), Hjärnfonden, Sweden (#FO2017-0243), the Swedish state under the agreement between the Swedish government and the County Councils, the ALF-agreement (#ALFGBG-715986), and European Union Joint Program for Neurodegenerative Disorders (JPND2019-466-236). KB has served as a consultant, at advisory boards, or at data monitoring committees for Abcam, Axon, Biogen, JOMDD/Shimadzu. Julius Clinical, Lilly, MagQu, Novartis, Roche Diagnostics, and Siemens Healthineers, and is a co-founder of Brain Biomarker Solutions in Gothenburg AB (BBS), which is a part of the GU Ventures Incubator Program (outside submitted work).MS is supported by the Knut and Alice Wallenberg Foundation (Wallenberg Centre for Molecular and Translational Medicine; KAW 2014.0363), the Swedish Research Council (#2017-02869), the Swedish state under the agreement between the Swedish government and the County Councils, the ALF-agreement (#ALFGBG-813971), and the Swedish Alzheimer Foundation (#AF-740191). MS has served on a scientific advisory board for Servier Pharmaceuticals (outside submitted work). AM, MJG, NJA, TKK, JLR, AS, and MSC report no competing interests. Data collection and sharing for this project was funded by the Alzheimer’s Disease Neuroimaging Initiative (ADNI) (National Institutes of Health Grant U01 AG024904) and DOD ADNI (Department of Defense award number W81XWH-12-2-0012). ADNI is funded by the National Institute on Aging, the National Institute of Biomedical Imaging and Bioengineering, and through generous contributions from the following: AbbVie, Alzheimer’s Association; Alzheimer’s Drug Discovery Foundation; Araclon Biotech; BioClinica, Inc.; Biogen; Bristol-Myers Squibb Company; CereSpir, Inc.; Cogstate; Eisai Inc.; Elan Pharmaceuticals, Inc.; Eli Lilly and Company; EuroImmun; F. Hoffmann-La Roche Ltd and its affiliated company Genentech, Inc.; Fujirebio; GE Healthcare; IXICO Ltd.; Janssen Alzheimer Immunotherapy Research & Development, LLC.; Johnson & Johnson Pharmaceutical Research & Development LLC.; Lumosity; Lundbeck; Merck & Co., Inc.; Meso Scale Diagnostics, LLC.; NeuroRx Research; Neurotrack Technologies; Novartis Pharmaceuticals Corporation; Pfizer Inc.; Piramal Imaging; Servier; Takeda Pharmaceutical Company; and Transition Therapeutics. The Canadian Institutes of Health Research is providing funds to support ADNI clinical sites in Canada. Private sector contributions are facilitated by the Foundation for the National Institutes of Health (www.fnih.org). The grantee organization is the Northern California Institute for Research and Education, and the study is coordinated by the Alzheimer’s Therapeutic Research Institute at the University of Southern California. ADNI data are disseminated by the Laboratory for Neuro Imaging at the University of Southern California.

## Supplementary material

Supplementary Figure 1.

Supplementary Figure 2.

Supplementary Figure 3.

